# *De novo* variants in *SNAP25* cause an early-onset developmental and epileptic encephalopathy

**DOI:** 10.1101/2020.05.11.20095141

**Authors:** Chiara Klöckner, Heinrich Sticht, Pia Zacher, Bernt Popp, Dewi P. Bakker, Katy Barwick, Michaela V. Bonfert, Eva H. Brilstra, Care4Rare Canada Consortium, Wendy K. Chung, Angus J. Clarke, Patrick Devine, Jennifer Friedman, Alyssa Gates, Gabriella Horvath, Jennifer Keller-Ramey, Boris Keren, Manju A. Kurian, Virgina Lee, Kathleen A. Leppig, Johan Lundgren, Marie T. McDonald, Amy McTague, Heather C. Mefford, Cyril Mignot, Mohamad A. Mikati, Caroline Nava, F. Lucy Raymond, Julian R. Sampson, Alba Sanchis-Juan, Vandana Shashi, Joseph T.C. Shieh, Marwan Shinawi, Anne Slavotinek, Tommy Stödberg, Nicholas Stong, Jennifer A. Sullivan, Ashley C. Taylor, Tomi L. Toler, Marie-José van den Bogaard, Saskia N. van der Crabben, Koen van Gassen, Richard H. van Jaarsveld, Jessica Van Ziffle, Alexandrea F. Wadley, Matias Wagner, Saskia B. Wortmann, Rikke S. Møller, Johannes R. Lemke, Konrad Platzer

## Abstract

**Purpose:** This study aims to provide the first comprehensive description of the phenotypic and genotypic spectrum of *SNAP25* developmental and epileptic encephalopathy *(SNAP25-* DEE) by reviewing newly identified and previously reported individuals.

**Methods:** Individuals harboring heterozygous missense or truncating variants in *SNAP25* were assembled through collaboration with international colleagues, matchmaking platforms and literature review. For each individual, detailed phenotyping, classification and structural modeling of the identified variant was performed.

**Results:** The cohort comprises 20 individuals with (likely) pathogenic de novo variants in *SNAP25*. Intellectual disability and early-onset epilepsy were identified as the core symptoms of *SNAP25*-DEE, with recurrent findings of movement disorders, cortical visual impairment and brain atrophy. Structural modeling for all variants predicted possible functional defects concerning SNAP25 or impaired interaction with other components of the SNARE complex.

**Conclusion:** We provide a first comprehensive description of *SNAP25*-DEE with intellectual disability and early onset epilepsy mostly occurring before the age of two years. These core symptoms and additional recurrent phenotypes show an overlap to genes encoding other components or associated proteins of the SNARE complex such as *STX1B, STXBP1* or *VAMP2*. Thus, these findings advance the concept of a group of neurodevelopmental disorders that may be termed “SNAREopathies”.

## INTRODUCTION

The neuronal SNARE complex (soluble N-ethylmaleimide-sensitive-factor attachment receptor complex) plays a central role in the regulation of synaptic signaling by mediating membrane docking, priming and fusion of synaptic vesicles with presynaptic membranes. This ultimately leads to the release of neurotransmitters into the synaptic cleft.^1-3^ During neuronal development, it promotes neurite outgrowth and the maturing process of synapses.^4,5^

The core SNARE complex comprises a four-helix bundle consisting of two SNAP25 helices (encoded by *SNAP25)*, one syntaxin 1A helix (encoded by *STX1A)*, and one synaptobrevin 2 helix (encoded by *VAMP2)*.^1,6,7^

Pathogenic *de novo* variants disrupting SNARE proteins *(VAMP2*, MIM 618760) or SNARE complex associated proteins, such as *STXBP1* (MIM 612164), *STX1B* (MIM 616172) are a known cause for neurodevelopmental disorders consisting of an overlapping phenotype of developmental delay (DD), intellectual disability (ID) and epilepsy.^8-12^

*SNAP25* showed a significant enrichment for *de novo* variants in a cohort study of individuals with neurodevelopmental disorders with epilepsy.^13^ Six individuals with heterozygous variants in *SNAP25*, four of them of *de novo* origin, have been described in single case reports showing developmental delay, seizures and variable neurological symptoms.^14-18^

The aim of this study is to establish a first comprehensive description of the phenotypic spectrum of *SNAP25* developmental and epileptic encephalopathy (*SNAP25*-DEE). We report 16 individuals harboring *de novo* variants in *SNAP25* and review all four previously published individuals with *de novo* variants determined to be (likely) pathogenic. Through molecular modeling, we provide further insights into possible mechanisms through which the identified variants may disrupt SNAP25 and the SNARE complex.

## MATERIALS AND METHODS

### Research Cohort and Identification of Variants

By using matchmaking platforms,^19^ personal communication with colleagues and a literature review, 27 individuals harboring heterozygous missense or truncating variants in *SNAP25* were assessed. After thorough evaluation, we included 20 individuals harboring *de novo* variants determined to be (likely) pathogenic, including 16 previously unreported individuals. The remaining seven individuals harbored variants of unknown significance (VUS, Supplement Tables S2, S3.2 and S4.3) and were not included in the phenotypic description.

Phenotypic and genotypic information were obtained from the referring collaborators by using a standardized questionnaire to evaluate clinical, electroencephalography (EEG), and cranial magnetic resonance imaging (cMRI) findings as well as variant information. Variants were identified using trio exome sequencing (ES), trio genome sequencing (GS) or gene panel sequencing. All examined individuals or their legal guardians provided written consent for clinical testing and publication according to the respective national ethics guidelines and with approval of the local ethics committees in the participating study centers.

According to data from gnomAD, *SNAP25* (NM_130811.4) shows a reduced number of truncating and missense variants in controls: (1) a pLI score = 0.99 with one truncating variant deposited at amino acid residue 204, two codons before the canonical stop and (2) a z-score for missense variants = 2.96. This indicates a selective constraint on these variant types in a control population not affected by early-onset NDD.^20^ Therefore, causality of both truncating and missense variants was assessed according to the guidelines of the American College of Medical Genetics (ACMG),^21^ focusing on the following criteria: PS2 *(de novo* origin), PM2 (absent in population databases), PP3 (multiple lines of computational evidence support a deleterious effect on the gene/gene product). For *in silico* prediction of missense variants, the following tools were used: CADD, REVEL, MutationTaster, M-CAP, Polyphen-2, GERP++. ^22-27^

### Structural Modeling

The structural effects of SNAP25 variants were modeled with SwissModel (Version 4.1.0)^28^ based on the experimental SNARE-αSNAP complex structures available (PDB codes: 3J96, 3J97, 3J98, 3J99, 6IP1, 6MDN).^29-31^ RasMol (Version 2.7.5)^32^ was used for structure analysis and visualization.

## RESULTS

The key clinical findings in *SNAP25*-DEE comprise DD and/or ID, early-onset seizures and variable neurological symptoms such as muscular hypotonia, movement disorders (ataxia, dystonia or tremor), cortical visual impairment (CVI) and brain volume loss. An overview of the main clinical symptoms is presented in Table 1.

**Table 1.** Summary of clinical findings in individuals with (likely) pathogenic *de novo* variants in *SNAP25*. Variant nomenclature corresponds to GenBank: NM_130811.4. Abbreviations: +, phenotype present (not specified); -, phenotype not present; ASD, autism spectrum disorder; AT, ataxia; CVI, cortical visual impairment; DD, developmental delay; ES, epileptic spasms; F, female; FS, focal seizures; GS, generalized seizures; ID, intellectual disability; M, male; m, months; MH, muscular hypotonia; n/a, not available; w, weeks; y, years.

| Individual | *de novo* variant | p. | Sex | Age | DD / ID | Seizures | Movement disorder | Neurological findings | cMRI | Other |
| --- | --- | --- | --- | --- | --- | --- | --- | --- | --- | --- |
| 1<sup>13</sup> | c.118A>G | p.(Lys40Glu) | M | 13y 2m | severe | GS | stereotypic hand movements | severe MH, CVI | normal | ASD, bilateral talipes, epicanthus, abnormal dentition |
| 2 | c.127G>C | p.(Gly43Arg) | M | 19y 5m | moderate | + | ataxia, tremor | MH, dystonia | normal | high arched palate, epicanthus |
| 3<sup>13</sup> |  |  | F | n/a | mild | GS | stereotypic hand movements, ataxia, |  | normal |  |
| 4 |  |  | F | 4y 3m | mild | GS | ataxia |  | normal |  |
| 5<sup>14</sup> | c.142G>T | p.(Val48Phe) | F | 20y | severe | GS, FS | ataxia | severe MH, dystonia | normal | high arched palate, hip dysplasia, joint hypermobility |
| 6<sup>16</sup> | c.149T>C | p.(Leu50Ser) | F | 5y 10m | + | ES |  |  | normal |  |
| 7 | c.170T>G | p.(Leu57Arg) | F | 0y 6w | profound | GS, FS |  |  | normal | deceased aged 40 weeks |
| 8<sup>15</sup> | c.200T>A | p.(Ile67Asn) | F | 11y | moderate | - | ataxia | MH | normal | eyelid ptosis |
| 9 | c.212T>C | p.(Met71Thr) | M | 8y 3m | moderate | - |  | MH | normal | ASD, bilateral talipes, epicanthus |
| 10 | c.212T>C | p.(Met71Thr) | M | 4y 11m | severe | GS, FS |  |  | normal |  |
| 11<sup>17</sup> | c.496G>T | p.(Asp166Tyr) | M | 23y | moderate | GS, FS |  |  | brain volume loss |  |
| 12 | c.497A>G | p.(Asp166Gly) | M | 17y | mild | GS, FS |  |  | normal |  |
| 13 | c.521A>C | p.(Gln174Pro) | M | 3y 6m | profound | ES, GS |  | MH, spasticity,<br>CVI | leukoencephalopathy,<br>brain volume loss | plagiocephaly |
| 14 | c.575T>C | p.(Ile192Thr) | F | 14y | moderate | ES | - |  | brain volume loss | macrocephaly, high arched palate |
| 15 | c.593G>C | p.(Arg198Pro) | F | 0y 3m | severe | ES |  | MH, spasticity,<br>CVI | normal |  |
| 16 | c.596C>G | p.(Ala199Gly) | F | 16y | severe | - | stereotypic hand<br>movements | CVI | leukoencephalopathy | plagiocephaly, hip dysplasia, joint<br>hypermobility, short stature |
| 17 | c.596C>T | p.(Ala199Val) | M | 3y 10m | moderate | GS, FS | stereotypic hand<br>movements, ataxia | MH | normal | ASD |
| 18 | c.114+2T>G | p.(?) | M | 2y 6m | mild | GS | ataxia | MH, dystonia | normal |  |
| 19 | c.520C>T | p.(Gln174*) | F | 2y | Profound | ES |  | severe MH,<br>spasticity, CVI | leukoencephalopathy,<br>brain volume loss | high arched palate |
| 20<sup>13</sup> |  |  | F | 0y 11m | + | GS |  |  | normal |  |

### Phenotypic spectrum

#### Global developmental delay / intellectual disability

All individuals presented with DD and a variable degree of ID, ranging between profound (3/18; 17%), severe (5/18; 28%), moderate (6/18; 33%), and mild (4/18; 22%).

Six individuals aged between 2 and 20 years remained non-verbal (6/14; 43%), with three being adolescent or adult. If language was acquired, individuals could either speak single words (2/14; 14%) or in sentences (6/14; 43%) with articulation noted to be poor or imprecise.

All individuals showed variable degrees of motor delay. Three individuals (3/14; 21%) aged older than three years were not able to walk, while three (3/14; 21%) needed assistance and seven individuals (7/14; 50%) were able to walk on their own.

Regression was reported in five individuals (5/14; 35%) with three of them showing signs of regression with the onset of seizures. This regression was primarily described as a loss of words previously learned.

#### Seizures

Seizures were reported in 17 individuals (17/20; 85%), while three individuals aged 8 to 16 years had no history of seizures. The age of seizure onset ranged between the 7th day of life to 12 years, with a median age of 12 months. In all but three individuals, seizures started before age two years. A broad spectrum including epileptic spasms, generalized and focal seizures were reported with most individuals showing multiple seizure types over time. Primarily generalized or focal to bilateral tonic-clonic seizures as well as epileptic spasms were the most common seizure type having occurred in five individuals (5/17; 29%). Additional seizure types include myoclonic, absence-like, tonic and atonic seizures, each reported for four individuals (4/17; 19%). Seizures reported in early childhood appeared to be more generalized in onset while older individuals aged 17 to 23 years predominantly exhibited (multi)focal epilepsies corresponding with respective EEG findings of multifocal epileptic discharges and generalized spike wave discharges. Seizure frequency ranged from numerous daily events (8/12; 67%) to sporadic (2/12; 11%). Response to antiepileptic drugs (AEDs) was inconsistent for individuals with recurrent seizures: seven of fourteen individuals (7/14; 50%), were treated with more than three AEDs and still had frequent seizures.

#### Brain MRI findings

cMRI was performed on 17 individuals and was unremarkable in eleven individuals (11/17; 65%). Focal or generalized brain volume loss was noted in four individuals (4/17; 24%) aged 7 months to 23 years and signs of a leukoencephalopathy were present in two individuals (2/17; 12%).

#### Neurological findings

The most common neurological finding was muscular hypotonia (10/15; 66%), with severe hypotonia leading to feeding difficulties being observed in four individuals (4/15; 27%). Spasticity was noted in three individuals (3/16; 19%). Further recurrent findings include movement disorder such as ataxia (7/16; 44%), dystonia (3/16; 19%) and tremor (2/16; 13%). CVI was described in five individuals (5/16; 31%).

#### Behavior

Most individuals were reported to have no behavioral issues. However, three individuals showed signs of an autistic spectrum disorder (3/16; 19%) and four individuals presented with repetitive mannerisms such as hand-flapping (4/16; 25%).

#### Additional findings

Musculoskeletal findings include bilateral clubfeet (3/17; 18%), hip dysplasia, and joint hypermobility, each being reported for two individuals (2/17; 12%). Most individuals had no or only minor dysmorphic features, with epicanthus being reported for three individuals (3/17; 18%). Further findings included a high-arched palate (4/17; 24%) with abnormal dentition or dental crowding (2/17; 12%; Supplemental Figure S1 shows recurrent combinations of symptoms and Supplemental Table S1 contains details on all phenotypic findings).

### Variant analysis

Out of the 20 enrolled individuals with *de novo* variants, 14 different missense variants were identified, in addition to two truncating variants (one nonsense variant and one splice donor variant).

All 16 variants were absent from the gnomAD database (last accessed April 2020). All (likely) pathogenic missense variants affected highly conserved amino acid residues (mean GERP++: 5.88) of the t-SNARE coiled-coil homology domain 1 (amino acid position 14-81) and t-SNARE coiled-coil homology domain 2 (amino acid position 135-202; Figure 1).^33^ All *de novo* missense variants were predicted to be deleterious by multiple *in silico* prediction programs (mean CADD: 30.1; for a complete overview of *in silico* analysis see Supplemental Tables S4.1 and S4.2).

Recurrent variants comprise the missense variant p.(Gly43Arg) identified in three individuals, the missense variant p.(Met71Thr) identified in two individuals, and the nonsense variant p.(Gln174^*^) identified in two individuals. Different missense variants affecting the same amino acid residue were also observed including p.(Asp166Gly) and p.(Asp166Tyr) as well as p.(Ala199Gly) and p.(Ala199Val).

**Fig. 1:**
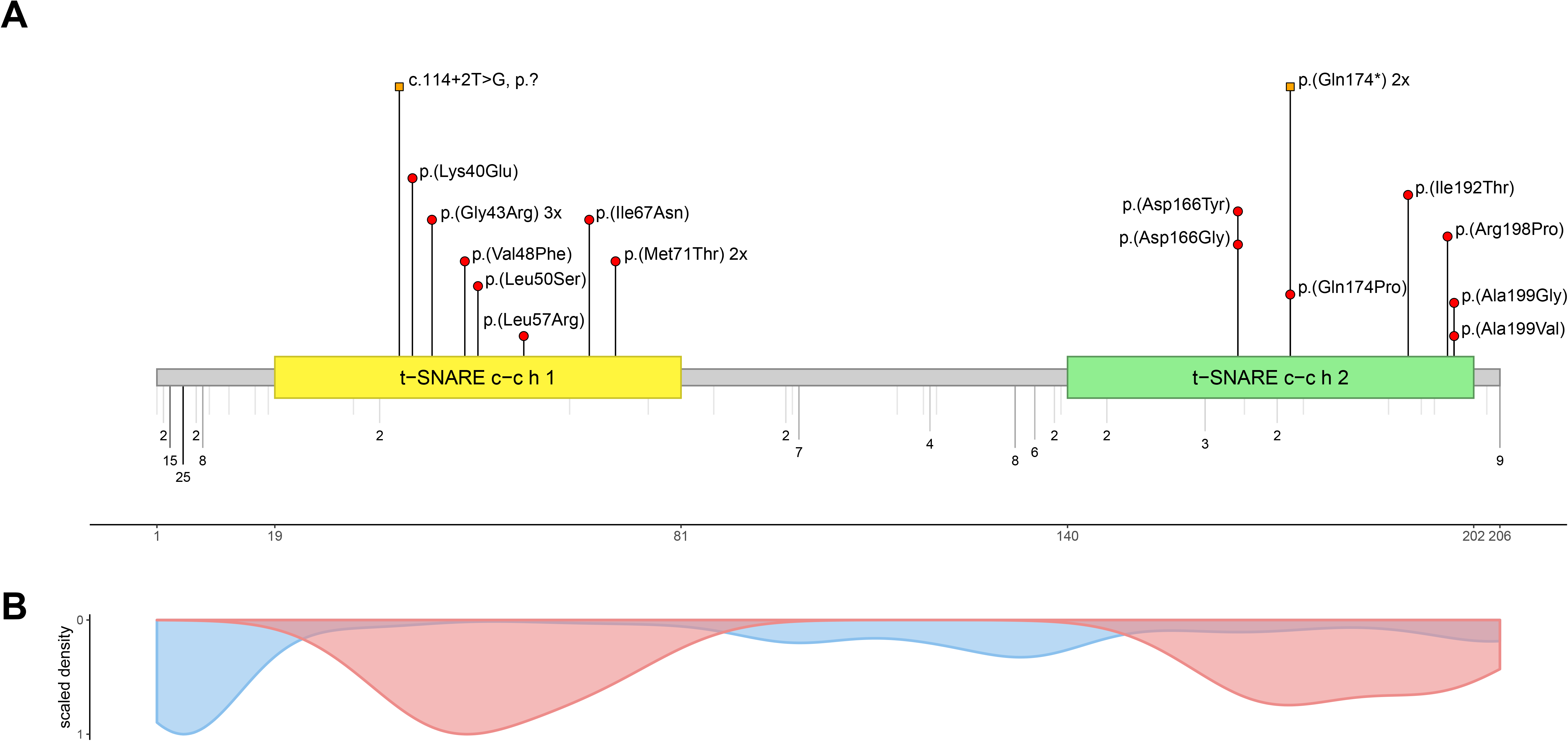
Overview of *SNAP25* variants (A) Location of missense and truncating variants in *SNAP25* with respect to domain structure (GenBank: NM_130811.4). Variants above protein scheme are *de novo* variants reported in this cohort with red circles indicating missense variants and orange squares indicating truncating variants. Below the protein scheme are missense variants in gnomAD with allele count and the degree of coloring being proportionate to the allele count with the lightest grey indicating singletons. Abbreviations are as follows: t-SNARE c-c h 1= t-SNARE coiled-coil homology domain 1; t-SNARE c-c h 2 = t-SNARE coiled-coil homology domain 2. (B) Density plot of all missense variants *(de novo* (likely) pathogenic variants in red and variants present in gnomAD in blue).

### *In silico* structural modeling

SNAP25 is part of the neuronal SNARE complex. The structure of this complex is shown in Fig. 2_AB indicating the positions of the variants detected in the present study. The sidechains of most residues affected are oriented towards the core of the SNARE complex, whereas few are oriented to the outside and interact with αSNAP, a protein that is involved in both SNARE assembly and disassembly after the completion of synaptic vesicle exocytosis.^34^

**Fig. 2:**
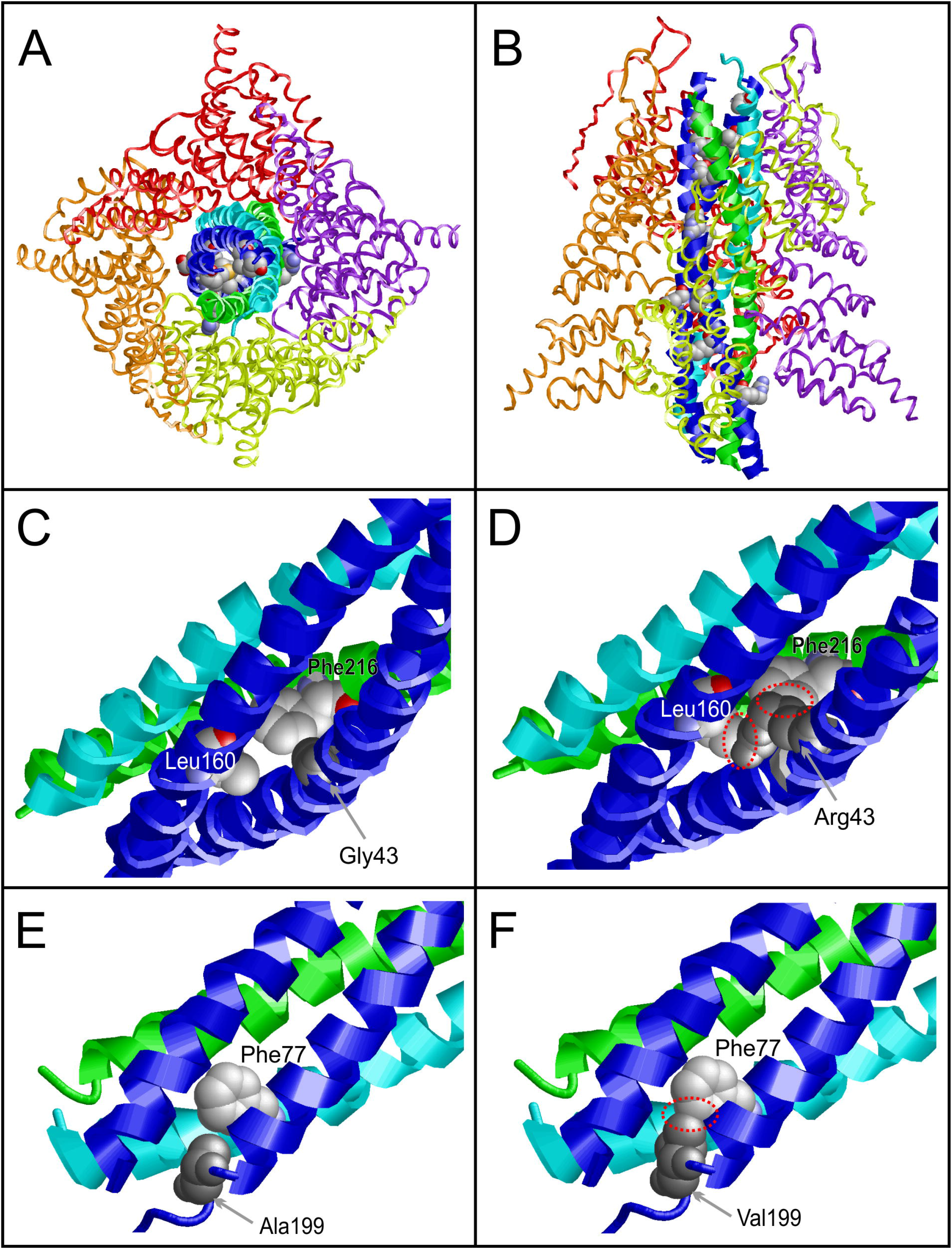
Location and structural effects of the SNAP25 variants detected in this study. (A) Top view of the SNARE-αSNAP complex. The individual proteins are shown in ribbon presentation and in different colors: SNAP25 (blue), Syntaxin (green), VAMP2 (cyan), αSNAP (yellow, orange, red, purple). Residues, for which variants were detected, are shown in space-filled presentation and colored according to their atom type (cpk coloring). (B) Side view of the complex shown in (A). (C) Interactions of Gly43 in the wildtype. Gly43 (gray) is located at a sterically demanding position of the four-helix bundle close to Leu160 of SNAP25 and Phe216 of syntaxin. (D) The longer sidechain of the p.(Gly43Arg) variant forms steric clashes (red dotted circles) with the Leu160 and Phe216 sidechain thereby destabilizing the helix bundle. (E) Interactions of Ala199 in the wildtype. Ala199 (gray) is located in spatial proximity to Phe77 of VAMP2. (F) The longer sidechain of the p.(Ala199Val) variant forms steric clashes (red dotted circle) with the Phe77 sidechain thereby destabilizing the SNAP25-VAMP2 interaction.

In order to better understand the effects of the identified variants, molecular modeling was performed. This analysis indicated that all (likely) pathogenic variants were predicted to destabilize the structure of SNAP25 by causing steric clashes: p.(Gly43Arg), p.(Leu57Arg), p.(Gln174Pro), by weakening intramolecular interactions: p.(Leu50Ser), p.(Lys40Glu), p.(Ile192Tyr), or by enhancing backbone flexibility: p.(Asp166Gly), p.(Ala199Gly). These structural effects are listed in more detail in the Supplemental Table S3.1. Other variants were predicted to predominantly destabilize the interactions with other components of the SNARE complex, namely STX1A: p.(Ile67Asn), p.(Met71Thr) or VAMP2: p.(Ala199Val). A third group of variants were predicted to result in disturbed interactions with αSNAP by causing steric clashes: p.(Val48Phe), p.(Asp166Tyr). It is important to note that some variants may both disturb SNAP25 structure and interactions, e.g. p.(Gly43Arg) or p.(Met71Thr). The structural effects of the p.(Gly43Arg) and p.(Ala199Val) variants are shown in detail in Fig. 2_CDEF. Taken together, despite differences in the proposed effects, all variants are expected to destabilize the SNARE complex itself or to affect its interactions with αSNAP (Supplemental Table S3.1).

## DISCUSSION

We present the first sizeable cohort of individuals with (likely) pathogenic *de novo* variants in *SNAP25* and provide a comprehensive evaluation of an early onset developmental and epileptic encephalopathy.

Moderate to profound DD and/or ID and early-onset seizures were identified as the core symptoms of *SNAP25*-DEE. In addition, ataxia, dystonia, CVI, brain volume loss, muscular hypotonia and spasticity were identified as recurrent clinical symptoms. Individuals with the most severe course of *SNAP25*-DEE exhibited profound DD, onset of seizures in the first two months of life, spasticity, CVI, and brain volume loss in three individuals.

Seizure semiology with a differentiation of focal or generalized onset can guide treatment options in individuals with epilepsy. An in-depth age-specific evaluation of seizure semiology was not possible in this comparatively small cohort, yet the adult individuals 5 and 11 exhibited mainly (multi)focal epilepsies. Video EEG data of individual 5 aged 20 years documented a focal epilepsy with parasagittal epileptic discharges corresponding with focal motor seizures (for the full video EEG report see Supplemental Table 1). This could indicate a possible evolution of seizures towards focal to bilateral tonic-clonic seizures in older individuals, but this will require further analysis of age-specific data on seizure semiology as well as their treatment to further elucidate *SNAP25*-DEE.

Of special interest for future clinical predictions is comparing the clinical course of individuals with recurrent variants or variants affecting the same amino acid residue: (1) The *de novo* missense variant p.(Gly43Arg) was identified in three individuals. It is remarkable that all individuals showed a rather mild phenotype comprising mild to moderate ID, sporadic generalized seizures that did not require treatment (individual 4) or showed good response to AEDs (individual 2) as well as ataxia and tremor. These finding suggests that this specific disruption of amino acid residue 43 could present with a milder variant-specific clinical course within *SNAP25*-DEE, while other individuals with (likely) pathogenic variants nearby in residues 40 and 48 presented with a more severe clinical course. (2) Two *de novo* variants affecting the amino acid residue 166, p.(Asp166Gly) and p.(Asp166Tyr) were identified in individual 11 and 12 aged 17 and 23 years, respectively. They presented with mild to moderate ID, focal and generalized seizures and did not show additional neurological symptoms. (3) A different picture is seen for the recurrent *de novo* variant p.(Met71Thr) that was identified in individual 9 and 10. Individual 9 was able to speak in sentences at the age of eight years and had no history of seizures while individual 10 was only able to speak single words at age seven years and had daily seizures since the age of two years and six months.

All (likely) pathogenic missense variants are located in the coiled-coil homology domains 1 and 2 of *SNAP25*, representing potential hotspots, also considering that little variation is observed in population databases in these domains (see Figure 1). The variants of four individuals aged two to 23 years with notable brain volume loss or signs of a leukoencephalopathy were located in the t-SNARE coiled-coil homology domain 2 (amino acid position 140-202), indicating a possible location-specific clinical observation (4/7; 57% of individuals with variants in this domain).

Future studies with additional individuals with *SNAP25*-DEE will shed more light onto the underlying clinical course. Recent modeling suggests an incidence of *de novo* variants in *SNAP25* in live births of approximately 1:100.000 for missense variants and 0.1:100.000 for nonsense variants.^35^

Data on functional analyses of variants in SNAP25 is scarce as of now as only an inhibition of synaptic vesicle exocytosis in a dominant negative manner has been confirmed previously for p.(Ile67Asn) with *in vitro* analysis of neuromuscular transmission.^15^ In the absence of comprehensive functional analyses, structural modeling of missense variants using published crystal structures can help in predicting the underlying mechanisms by which variants disrupt either SNAP25 or its interaction with other proteins of the SNARE complex.

In some instances, individuals harboring variants for which similar structural changes were predicted showed a similar course of disease. (1) The variants identified in individuals 9, 10 (p.(Met71Thr)) and 14 (p.(Ile192Thr)) are both predicted to cause a reduced packing with each other. All three individuals presented with moderate to severe ID while motor development seemed to be only mildly affected with all of them being able to walk on their own. In addition, individuals 10 and 14 presented with a rather late onset of seizures at 25 years and 12 years respectively, while individual 9 did not have a history of seizures. (2) Structural modeling of the likely pathogenic variants p.(Gln174Pro) and p.(Arg198Pro) indicated a disruption of the helix of SNAP25 for both variants. Individuals 13 (p.(Gln174Pro)) and 15 (p.(Arg198Pro)) presented with a severe course of disease with a seizure onset at age 2 months, severe to profound DD, CVI and spasticity. (3) Of further interest are two variants affecting the amino acid residue 199, p.(Ala199Gly) (individual 16) and p.(Ala199Val) (individual 17) with regards to a possible interaction with the SNARE complex protein VAMP2. The variant p.(Ala199Val) is predicted to cause a steric clash with the amino acid residue 77 of VAMP2. In a recent study, three individuals with variants affecting the amino acid positions 75, 77 and 78 of VAMP2 were described.^9^ All individuals showed overlapping clinical symptoms to the two individuals of the current *SNAP25* cohort comprising moderate to severe ID, onset of seizures in the first year of life, muscular hypotonia, ASD, stereotypic hand movements, CVI, absent speech and unremarkable brain imaging.^9^ These findings indicate that the disruption of these structural domains in either SNAP25 or VAMP2 may cause a similar downstream functional effect and result in a similar clinical course. Putting these clinical observations in *SNAP25*-DEE in context to other known disease genes of the neuronal SNARE complex, it becomes clear that a shared clinical course has been repeatedly described, suggesting a shared phenotypic spectrum of the neuronal “SNAREopathies” (see Supplemental Table S5 and Figure S2)^9,12,36^

The data on truncating variants including the recurrent *de novo* nonsense variant in two individuals and the *de novo* splice site variant in one individual is not as complete due to the relatively small cohort assembled. The nonsense variant p.(Gln174*) is located 32 base pairs from the end of exon seven of eight, so nonsense mediated mRNA decay (NMD) cannot be readily assumed, and the variant more likely leads to the translation of a truncated protein.^37^ Nonetheless, supporting causality of the nonsense variant p.(Gln174*) is the fact that it is predicted to truncate SNAP25 by 33 amino acids in a highly conserved region and the occurrence of other (likely) pathogenic missense downstream of position 174. The other potentially truncating splice site variant c.114+2T>G, p.(?) is located at the donor site of exon three. This variant could lead to an in-frame exon skipping possibly resulting in a non-functional protein that is quickly dismantled (for a report on splice prediction see Supplemental Table S4.2). Exon three contains 14 amino acids until amino acid position 38 and all (likely) pathogenic missense variants in this cohort were located in downstream exons. The affected individual 18 presented with mild developmental delay, ataxia and frequent seizures fitting the spectrum of *SNAP25*-DEE. As the variant occurred *de novo*, it can be classified as likely pathogenic. In contrast, a mildly affected individual with mild ID and no seizures inherited the truncating variant c.464delG p.(Gly155Alafs*84) that was classified as a VUS from their unaffected mother (see individual V6, Supplemental Table 2). The variant also likely escapes NMD but due to the substantial change in the amino acid sequence possibly results in a non-functional protein that is quickly dismantled. In addition to *SNAP25* having a pLI-score of 0.99 and only one reported truncating variant at amino acid position 204 of 206 (p.Gly204^*^) in the gnomAD database, different mouse models support the potential role of haploinsufficiency in the origin of *SNAP25*-DEE, since SNAP25(+/-) mice display a susceptibility to induced seizures, EEG abnormalities and cognitive deficits.^38^ Haploinsufficiency of *SNAP25* may therefore lead to a rather mild phenotype but the preliminary findings reported in this cohort must be complemented by future analyses. Similar observations have been made concerning *STX1B*, where truncating variants cause mild ID with epilepsy whereas missense variants cause a more severe form of DEE.^36^ This phenomenon of truncating variants resulting in a rather mild course of disease is also known for multiple other NDD genes encoding ion channels, e.g. *GRIN2A* or KCNQ2.^39,40^

In summary, *de novo* variants in *SNAP25* cause an early onset developmental and epileptic encephalopathy mainly characterized by ID and epilepsy, demonstrating a strong phenotypic overlap with disorders caused by the disruption of other components or associated proteins of the SNARE complex, including movement disorders, cortical visual impairment and brain atrophy. These findings add to the delineation of a group of disorders that may be called “SNAREopathies”.

## Data Availability

All relevant data and methods are reported in the article and in the supplement.

## Acknowledgements

We thank the patients and their families for their participation and support of this study. We especially thank Elizabeth Dellureficio for her continuous efforts to bring affected families together.Work on individual 5 was supported in part by grants from SFARI and the JPB Foundation. We thank the clinicians involved with the care of these patients including Dr Stephen Nirmal, Dr Alasdair Parker and Dr Manali Chitre, UK. AM, KB and MAK are funded by the NIHR GOSH BRC. The views expressed are those of the author(s) and not necessarily those of the NHS, the NIHR or the Department of Health.

Individual 7 was enrolled in the NIHR BioResource research study, supported by the Cambridge Biomedical Research Centre and the National Institute for Health Research (NIHR) for the NIHR BioResource (grant number RG65966).

Individual 16 was ascertained in the Duke Genome Sequencing Clinic. Funding for the Duke Genome Sequencing Clinic which is supported by the Duke University Health System. Individual V6 was enrolled in Care4Rare Canada Consortium, funded by Genome Canada, Ontario Genomics Institute (OGI-147), Canadian Institutes of Health Research, Ontario Research Fund, Genome Alberta, Genome British Columbia, BC Children’s Hospital Foundation, BC Children’s Hospital Research Institute, BC Provincial Health Services Authority, Genome Quebec, and Children’s Hospital of Eastern Ontario Foundation.

This study makes use of data generated by the DECIPHER Consortium. A full list of centres who contributed to the generation of the data is available from https://decipher.sanger.ac.uk/ and via email from. Funding for the project was provided by the Wellcome Trust.

R.S.M. was supported by a grant from the Lundbeck Foundation (R277-2018-802).

Research reported in this publication was supported by the National Human Genome Research Institute of the National Institutes of Health under Award Number U01HG009599.

The content is solely the responsibility of the authors and does not necessarily represent the official views of the National Institutes of Health. Amendola et. al. American Journal of Human Genetics. 2018 Sep 6;103(3):319-327. doi: 10.1016/j.ajhg.2018.08.007

## Web Resources

DECIPHER, https://decipher.sanger.ac.uk/

gnomAD, http://gnomad.broadinstitute.org/

GeneMatcher, https://genematcher.org/

GenBank, https://www.ncbi.nlm.nih.gov/genbank/

OMIM, http://www.omim.org/

R, http://www.r-project.org/

UniProt database, https://www.uniprot.org/

## Supplemental Data

Supplemental Data includes seven Supplemental Tables and two Supplemental Figures.

